# Methylome-wide analysis reveals epigenetic marks associated with resistance to tuberculosis in HIV-infected individuals from East Africa

**DOI:** 10.1101/2020.07.14.20153395

**Authors:** Catherine M. Stein, Penelope Benchek, Jacquelaine Bartlett, Robert P. Igo, Rafal S. Sobota, Keith Chervenak, Harriet Mayanja-Kizza, C. Fordham von Reyn, Timothy Lahey, William S. Bush, W. Henry Boom, William K. Scott, Carmen Marsit, Giorgio Sirugo, Scott M. Williams

**Affiliations:** Department of Population and Quantitative Health Sciences, Case Western Reserve University, Cleveland, Ohio; Division of Infectious Disease and HIV Medicine, Department of Medicine, Case Western Reserve University, Cleveland, Ohio; Department of Genetics and Genome Sciences, Case Western Reserve University, Cleveland, Ohio; Department of Neurology, Feinberg School of Medicine, Northwestern University, Chicago, Illinois; Department of Medicine and Mulago Hospital, School of Medicine, Makerere University, Kampala, Uganda; Geisel School of Medicine at Dartmouth, Hanover, New Hampshire; John P. Hussman Institute for Human Genomics, University of Miami, Miami, Florida; Rollins School of Public Health, Emory University, Atlanta, Georgia; Department of Systems Pharmacology and Translational Therapeutics, University of Pennsylvania, Philadelphia, Pennsylvania

**Author notes:** These authors contributed equally as first authors of this work (Catherine M. Stein, Penelope Benchek). These authors contributed equally as senior authors of this work (Giorgio Sirugo, Scott M. Williams).

**Keywords:** methylation, epigenetics, infectious disease, genetics, genomics, lung function, immunology

## Abstract

**Background:** Tuberculosis (TB) is the most deadly infectious disease globally and highly prevalent in the developing world, especially sub-Saharan Africa. Even though a third of humans are exposed to *Myocbacterium tuberculosis* (Mtb), most infected immunocompetent individuals do not develop active TB. In contrast, for individuals infected with both TB and the human immunodeficiency virus (HIV), the risk of active disease is 10% or more per year. Previously, we identified in a genome-wide association study a region on chromosome 5 that was associated with resistance to TB. This region included epigenetic marks that could influence gene regulation so we hypothesized that HIV-infected individuals exposed to Mtb, who remain disease free, carry epigenetic changes that strongly protect them from active TB. To test this hypothesis, we conducted a methylome-wide study in HIV-infected, TB-exposed cohorts from Uganda and Tanzania.

**Results:** In 221 HIV-infected adults from Uganda and Tanzania, we identified 3 regions of interest that included markers that were differentially methylated between TB cases and LTBI controls, that also included methylation QTLs and associated SNPs: chromosome 1 (*RNF220*, p=4×10^−5^), chromosome 2 (between *COPS8* and *COL6A3* genes, p=2.7×10^−5^), and chromosome 5 (*CEP72*, p=1.3×10^−5^). These methylation results colocalized with associated SNPs, methylation QTLs, and methylation × SNP interaction effects. These markers were in regions with regulatory markers for cells involved in TB immunity and/or lung.

**Conclusion:** Epigenetic regulation is a potential biologic factor underlying resistance to TB in immunocompromised individuals that can act in conjunction with genetic variants.

## BACKGROUND

*Mycobacterium tuberculosis* (Mtb), the cause of tuberculosis (TB), results in approximately 1.5 million deaths per year [1], but the vast majority of the almost 2 billion people infected with Mtb do not progress to active TB. Although the risk of developing active TB is low in most infected people, it is the most common cause of death in people with HIV infection who live in TB endemic countries [2, 3]. As many as 10% of co-infected people develop active disease each year, illustrating how strongly immune compromise contributes to risk. People with HIV infection, who do not develop active TB despite Mtb infection, offer a major opportunity to understand resistance to TB that persists despite being immunocompromised, and possibly a key to how any Mtb infection leads to active TB.

Several studies indicate that susceptibility (or resistance) to TB is at least partially due to genomic factors [4]. Indeed, several genome-wide association studies (GWAS) have been conducted, identifying loci of interest, but most of these studies show small effect sizes in HIV uninfected subjects. In contrast, our GWAS [5], conducted in HIV-infected subjects, found significant association with resistance to TB and a region of chromosome 5 containing the *IL12B* gene. Annotation of this region showed that the associated SNPs resided in a histone mark, indicating that epigenetic marks in this region may influence regulation of a nearby gene that we hypothesized to be *IL12B*. Because differences in genomic features, such as histone and other epigenetic marks, can impact gene expression patterns, thereby linking genetic and environmental disease risk factors, and since epigenetic marks can be inherited, they have been hypothesized to explain some of the “missing heritability” for complex diseases [6-8]. Evidence for this model has been found in cancer and autoimmune diseases, where associations have been demonstrated between epigenetic marks and disease risk [9, 10]. Based on these studies and our own prior results on chromosome 5, we hypothesized that regulatory factors, including methylation marks, associate with TB susceptibility/resistance, beyond the genetic influences encoded in the DNA.

Since disease risk may be influenced by epigenetic variation, either directly or indirectly via DNA-level variation [11], we examined loci identified through an methylome-wide analysis (MWAS) and their association with single nucleotide polymorphisms (SNPs) at those same loci. We also investigated differential methylation near loci initially identified through GWAS of TB risk (Figure 1). Our underlying model was that both genetic and epigenetic factors are associated with risk of TB and that the two may modify the effects of each other. To understand the functional implications of identified loci, we annotated these loci using available databases. By integrating results across multiple platforms and data types [12], we extended GWAS results to show that a framework containing both epigenetic changes and DNA variations, and their interactions, can associate with regulation of lung and immune cell function relevant to TB pathogenesis.

**Figure 1.**
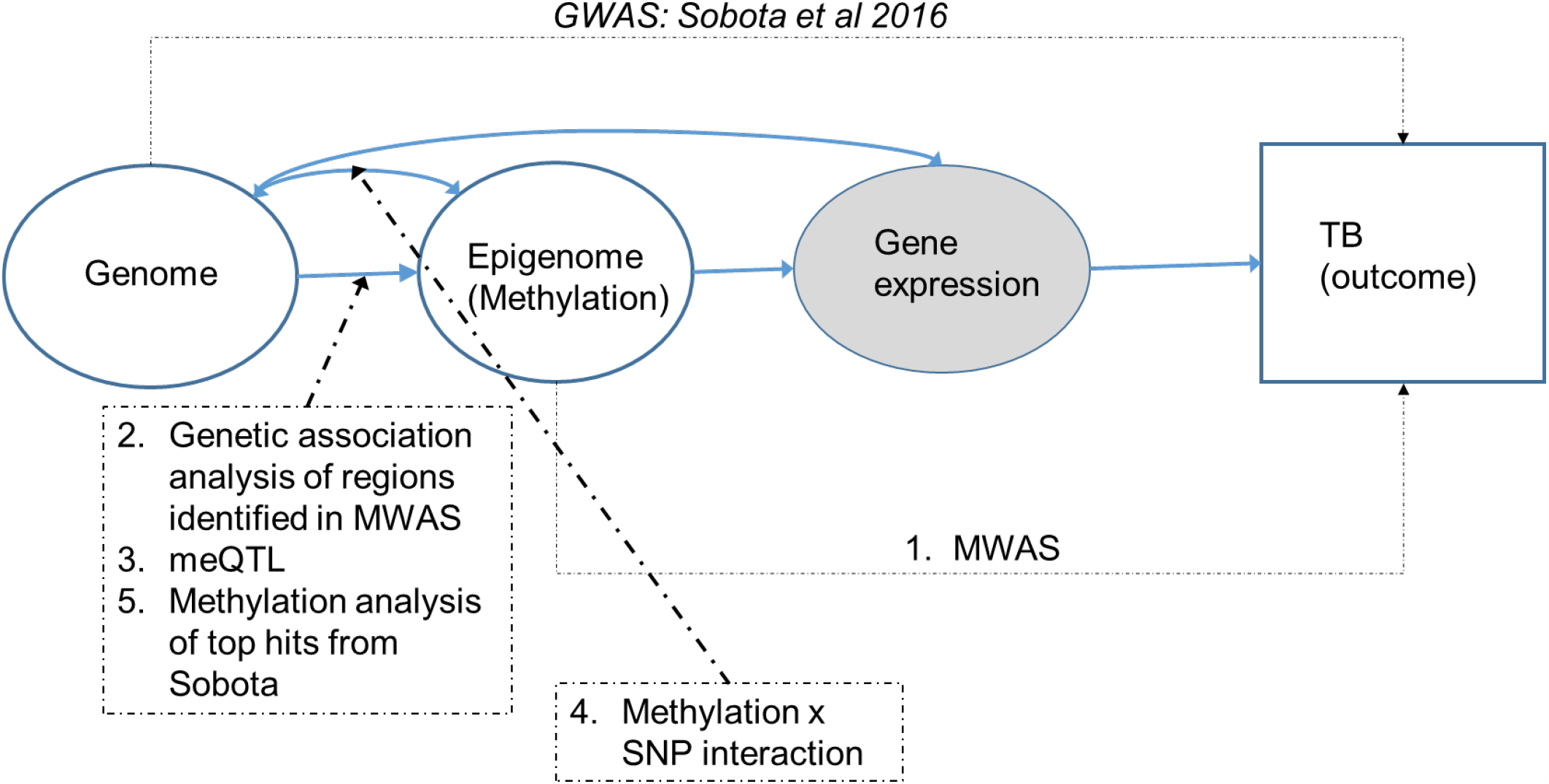
Analytical framework

## RESULTS

### Overall approach

Our approach (Figure 1) was to first conduct an MWAS in the Ugandan and Tanzanian cohorts independently. Because the Ugandan cohort was larger, we considered it the discovery cohort and the Tanzanian cohort as the replication set, although we repeated analyses in the opposite direction. Markers that were differentially methylated in the discovery cohort at p< 5×10^−5^ to account for multiple testing, and p<0.1 in the replication cohort were considered for subsequent analyses. In the next step, we examined SNPs from our GWAS ± 200 kb of the methylation signal, and identified whether they were significantly (p<0.05) associated with TB; we did not adjust for multiple testing because these and subsequent analyses were hypothesis-driven. In the third step, we conducted a meQTL analysis, where we examined SNPs ± 200 kb of the associated methylation marker to test for association with methylation of the associated CpG site. Fourth, we examined the interaction between methylation status and SNPs in association with TB. A comparison of the results of the second through fourth steps addressed whether genetic variation at least partially influences TB susceptibility through effects on methylation, or if differential methylation and genetic variation acted independently. Lastly, all loci significant in the first step underwent functional annotation as described in the Methods. In parallel, we examined the association between methylation level and TB in regions that were significantly associated with TB in our GWAS (Table 1 from Sobota et al.[5]).

**Table 1.**
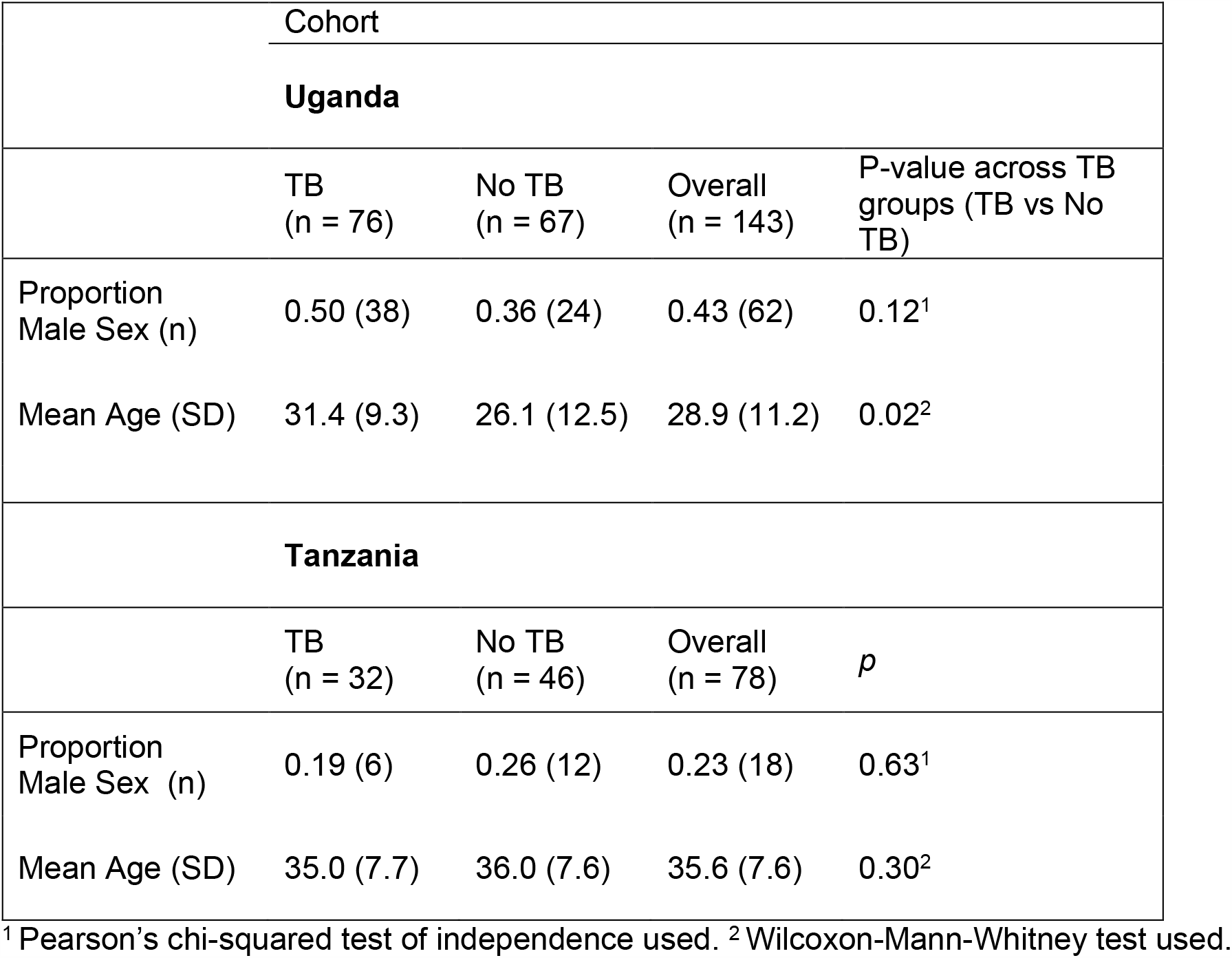
Study population characteristics

| Cohort |  |  |  |  |
| --- | --- | --- | --- | --- |
| Uganda |  |  |  |  |
|  | TB<br>(n = 76) | No TB<br>(n = 67) | Overall<br>(n = 143) | P-value across TB<br>groups (TB vs No<br>TB) |
| Proportion<br>Male Sex (n) | 0.50 (38) | 0.36 (24) | 0.43 (62) | 0.12<sup>1</sup> |
| Mean Age (SD) | 31.4 (9.3) | 26.1 (12.5) | 28.9 (11.2) | 0.02<sup>2</sup> |
| Tanzania |  |  |  |  |
|  | TB<br>(n = 32) | No TB<br>(n = 46) | Overall<br>(n = 78) | *p* |
| Proportion<br>Male Sex (n) | 0.19 (6) | 0.26 (12) | 0.23 (18) | 0.63<sup>1</sup> |
| Mean Age (SD) | 35.0 (7.7) | 36.0 (7.6) | 35.6 (7.6) | 0.30<sup>2</sup> |

<sup>1</sup> Pearson's chi-squared test of independence used. <sup>2</sup> Wilcoxon-Mann-Whitney test used.

### Sample description

This analysis included a total of 221 adult subjects with HIV infection (Table 1). The Uganda cohort included 143 subjects, of whom 76 had TB. The Tanzania cohort included 78 subjects, of whom 32 had TB. The subjects who did not have active TB all had latent Mtb infection (LTBI) based on a positive tuberculin skin test. No subjects received antiretroviral therapy (ART) for HIV because all were enrolled prior to the availability of ART in Africa. In the Uganda cohort, there was a greater proportion of males among TB cases than controls, reflecting the greater preponderance of TB among males in the general population. Thus, sex was considered as a covariate in all analyses.

### Differentially methylated regions associated with TB

In our analysis, we identified 3 regions, the first two on chromosomes 1 and 2, respectively, that were differentially methylated with p < 5 × 10^−5^ in the Uganda sample and p < 0.10 in the Tanzania cohort with effect estimates in the same direction (Table 2) and a third region, on chromosome 5 that showed a significant (p=2×10^−5^) differentially methylated marker detected in the Tanzanian cohort that replicated in Ugandan cohort (p=0.0398) and had effect estimates in the same direction in both populations. A nearby chromosome 5 marker also showed association in the Ugandan cohort (p=2.08×10^−5^), but did not replicate in the Tanzanian cohort. The two methylation markers on chromosome 5 were uncorrelated in both cohorts (r=0.03 in Uganda, r=0.18 in Tanzania). The marker on chromosome 1 fell in a methylation island in *RNF220* (Figure 2a), the marker on chromosome 2 fell in an “open sea” (CpG sites not associated with a CpG island or CGI) flanked by *COPS8* and *COL6A3* (Figure 2b), and the markers on chromosome 5 were on the “south shore” and “north shore” (regions up to 2 kb from a CGI), respectively, of the *CEP72* gene (Figure 2c).

**Table 2.** Significantly associated methylation markers

| Methylation marker name | chr | position | Relation to Island | UCSC_RefGene_Name | Uganda p-value | Tanzania p-value | Combined p-value |
| --- | --- | --- | --- | --- | --- | --- | --- |
| *Methylation markers that are replicated in both cohorts* |  |  |  |  |  |  |  |
| cg19382731 | chr1 | 44883990 | Island | RNF220 | 4.62 E-05 | 0.07 | 4.63E-05 |
| cg16974832 | chr2 | 238188707 | OpenSea | Flanking genes (COPS8 and COL6A3) | 4.47 E-05 | 0.04 | 2.69E-05 |
| *Different methylation markers in same region by population attaining significance* |  |  |  |  |  |  |  |
| cg18730862 | chr5 | 611926 | N_Shore | CEP72 | 2.08E-05 | 9.21E-01 | 2.28E-04 |
| cg03602880 | chr5 | 612961 | S_Shore | CEP72 | 0.0398 | 2.09E-05 | 1.25E-05 |

GWAS: Sobota et al 2016

**Figure 2.**
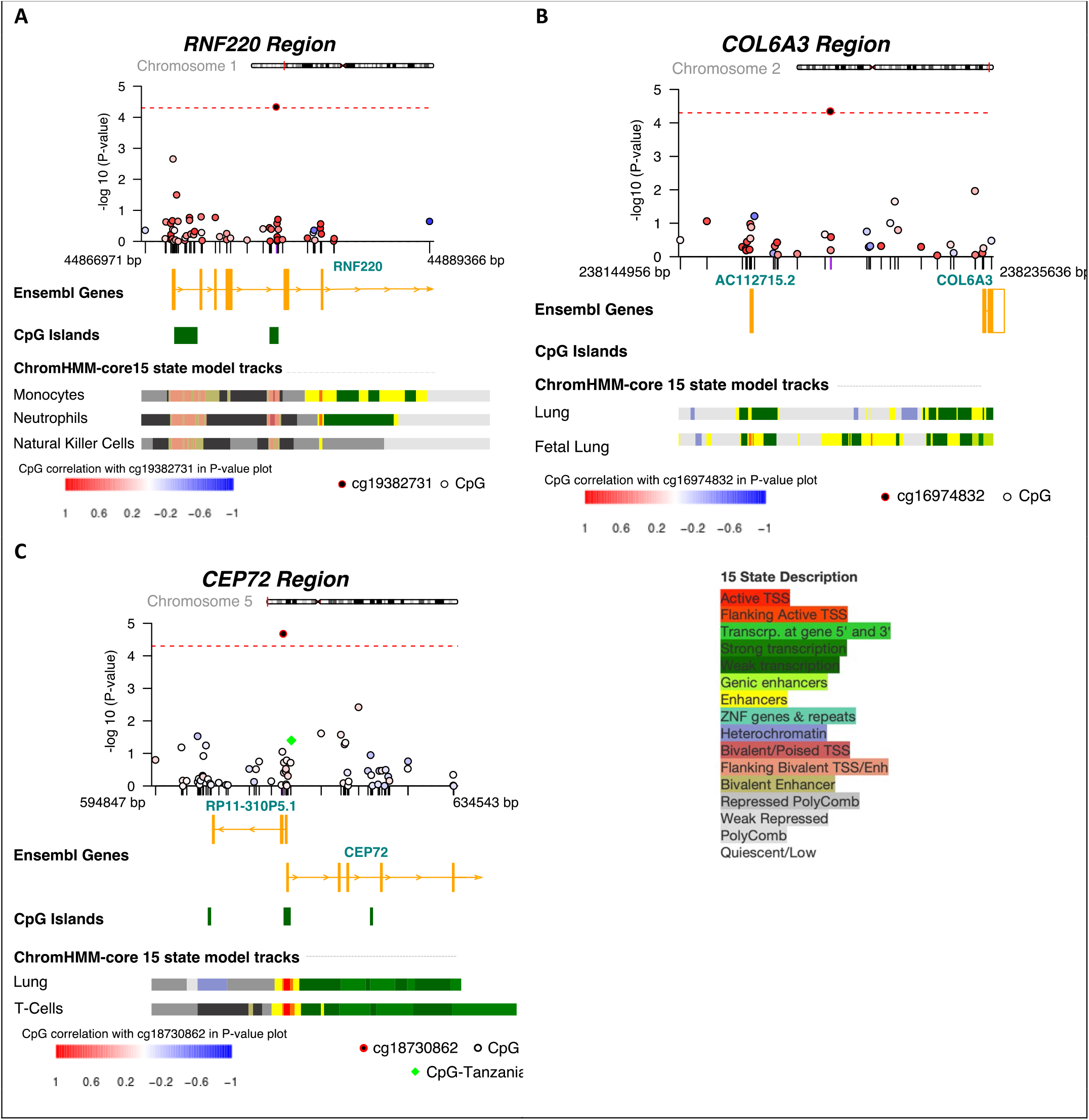
Regional plots of epigenetic-phenotype association results. CoMET plots of our top CpG methylation association results are shown. The top track shows the methylation-phenotype association P-values across the region for the Uganda cohort, with CpG sites depicted as circles. Also shown in the top track are the estimated co-methylation values (correlation in DNA methylation values) between the reference (top) CpG site and others in the region, depicted by the fill of the circle, as a gradient from red to blue (correlation mapping shown in legend at the bottom of each panel). Additional tracks include Ensembl genes, CpG islands and regulatory information for the specified tissues via the ChromHMM-core 15 state model. Ensembl genes shown may be truncated if they extend outside of viewing window. A) RNF220 gene CpG site cg1938271 shown +/-25kb. B) COL6A3 gene CpG site cg16974832 shown +/-50kb. C) CEP72 gene CpG site cg18730862 shown +/-25kb. The CpG-Tanzania indicated site is for cg03602880 (*p*=2×10^−5^ in the Tanzania cohort, *p*=0.04 in the Uganda cohort); Spearman’s correlation between cg030602880 and cg18730862 was 0.11 overall (0.03 in Uganda only and 0.18 in Tanzania only). D) Legend for panels A-C. The 15 chromatin states and corresponding color mappings.

### Validation of differentially methylated regions in GWAS

We then interrogated the regions on chromosomes 1, 2, and 5 that contained differentially methylated CpG sites in the GWAS dataset that included the same subjects [5] (Supplemental Table 1). All three regions had a SNP associated with TB within 200 kb of the differentially methylated sites (p<0.05 unadjusted for multiple testing): chromosome 1, rs175222 (p=0.00016) (Figure 3a), chromosome 2 rs7586225 (within *COL6A3*, p=0.0082) (Figure 3b), and chromosome 5, rs12518227 (p=0.018) (figure 3c). Associated SNPs in these regions are in regulatory regions, or in introns, and one on chromosome 5 is a missense variant (rs868649). One of these SNPs (rs175222 on chromosome 1) maintained significance after Bonferroni correction.

**Figure 3.**
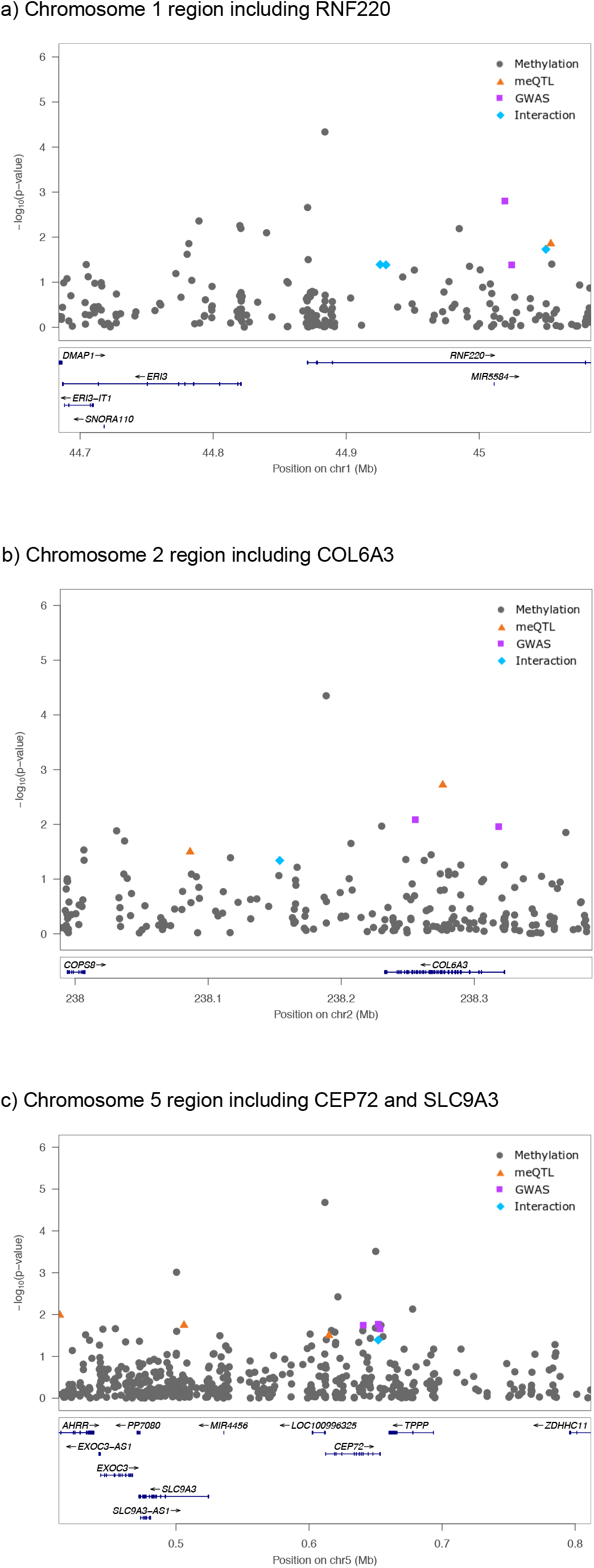
Plots of association p-values from MWAS, GWAS, meQTL, and methylation × SNP interaction analyses. MWAS p-values come from Uganda cohort, as it was the discovery cohort, and GWAS, meQTL, and methylation × SNP p-values come from the combined cohort. Results from each type of analysis are represented by different colors and shapes, with –log(p-value) plotted on the y-axis, and chromosomal location on the x-axis. a) chromosome 1 (20 SNPs), b) chromosome 2 (33 SNPs), c) chromosome 5 (22 SNPs)

### SNP-methylation QTL associations and SNP × methylation interaction

Next we examined both association between SNPs in these regions and the level of methylation of the methylation site (meQTL analysis), and also examined the interaction between SNP genotype and methylation marker in its association with TB. These results are shown in Figure 3 to illustrate their co-localization with the original methylation findings and marginal SNP associations with TB. Again, each region had a significant meQTL effect and SNP × methylation interaction effect. On chromosome 1 (Figure 3a), rs928685 was a significant meQTL (p=0.014), and there were 3 SNPs with significant interaction effects with the methylation site (rs270709, p=0.0108; rs6664827, p=0.0405; rs1890948, p=0.0412), all within the *RNF220* gene. On chromosome 2 (Figure 3b), there were 2 meQTLs — rs2645771, within *COL6A3* (p=0.0019), and rs10165956 (p=0.0318) — and 1 methylation x SNP interaction at rs4530312 (p=0.0457). On chromosome 5 (Figure 3c), there were 3 meQTLs: rs4956936 (within *AHRR*, p=0.0103), rs1697952 (p=0.0316), and rs6864158 (within *SLC9A3*, p=0.0392). This region had one SNP that interacted with methylation status, rs12518227 (within *CEP72*, p=0.0405). The SNPs that associate with methylation and those that associate with TB are in all cases different.

### Functional implications of 3 differentially methylated regions

The methylation site on chromosome 1 falls in a frequently interacting region (FIRE) and is a bivalent enhancer in a variety of cells involved in TB immunity, including T cells, monocytes, and B cells; bivalent enhancers have been linked to increased gene expression [13]. The chromosome 2 region contains a histone modifier, and is an enhancer in lung tissue; a FIRE crosses the *COL6A3* gene. The differentially methylated region on chromosome 5 falls within an active transcription start site in T cells and lung tissue, and flanks the transcription start site in monocytes, B cells and neutrophils. Tracks showing functional regions in these regions are shown in Figure 2.

### SNPs significant in original GWAS are also in differentially methylated regions

We also examined loci that were genome-wide significant in our prior GWAS for TB [5], and found that they contained significantly differentially methylated sites. The MWAS did not identify these regions as having significant methylation effects after multiple testing correction. However, we explored these based on our original hypothesis that regions associated with TB risk in GWAS might also act through epigenetic mechanisms. In most cases, the same methylation marker was not significant in both the Uganda and Tanzania datasets. For the methylation marks nearest *IL12B*, the most significant finding from our GWAS [5], differential methylation associated with TB in both Uganda (cg15353886, p=0.0015) and Tanzania (cg11092268, p=0.0089). Two other methylation markers that replicated in the two samples from these regions fell on chromosome 5 (cg09049927, p=0.00089 in combined data) and chromosome 17 (cg24357302, p=0.00071 in combined data). Both of these methylation markers fell in “open seas”, with the chromosome 5 marker falling near an miRNA, and the chromosome 17 in an intron of *ABCA8* (Supplemental Table 2).

## DISCUSSION

Host factors play an important role in the progression from LTBI to active TB disease, including genetic and transcriptomic factors [4, 14]. To our knowledge, only two small methylome-wide studies have been published [15, 16] in human cohorts, though *in vitro* studies have been conducted [17-19]. Our analysis of TB patient cohorts from Uganda and Tanzania revealed three regions that were differentially methylated in HIV-infected individuals who exhibited protection from active TB. These same regions contained nominally significant SNPs associated with TB, as well as SNPs associated with methylation level and SNPs interacting with methylation level in association with TB. Functional annotation revealed that these loci have regulatory effects on cells involved in the TB immune response, providing mechanistic plausibility of the results. In addition, regions that were previously shown to have genome-wide significant associations between SNPs and TB also demonstrated differential methylation. Although none of these analyses alone provide compelling evidence for association, the concordance of the different analyses does; these results indicate that epigenetic factors, in combination with genetic variation, can influence susceptibility to TB.

Cellular and humoral immunity are well established components of the immune response to Mtb, but our results newly establish that epigenetic regulation of T cell, B cell and monocytes can influence protection from active TB. This may define a path from SNP to regulation of gene expression to protection from TB disease as well as suggest new drug targets for prevention of active TB. As seen in Figure 2, the three regions identified through our analyses contain regulatory markers for monocytes, T cells, natural killer cells, and lung tissue, among others, suggesting a role for these specific loci in TB susceptibility.

While the methylation marker on chromosome 2 resides in an “open sea”, the SNPs associated with TB and methylation level, as well as SNP x methylation interaction, are all within *COL6A3*. Collagen VI, as a component of the extracellular matrix, plays a role in innate immune defense against bacteria and regulates autophagy [20, 21], thus indicating that COL6A3 influences protection from TB via influencing immune responses to TB antigens. Collagen VI-related myopathies are associated with decreased pulmonary function [22]. *COL6A3* has been associated with lung cancer [23]. Thus, we hypothesize that *COL6A3* may influence TB susceptibility through its effect on lung function. The fact that the methylation site we identified falls in an enhancer region for lung tissue reinforces this hypothesis. The mechanism of action may be diverse as the associated SNPs are also both regulatory and result in coding changes. *RNF220* is in the middle of the chromosome 1 region, and many of the associated SNPs are within the *RNF220* coding region (Figure 3a). RNF220 is known to enhance Wnt signaling [24], and thus may indicate a role for epigenetic modulation of Wnt signaling in the innate immune response to Mtb [25].

The associated methylation sites and SNPs on chromosome 5 cross multiple genes, but most results co-localize to *SLC9A3* (Figure 3c). Another methylome-wide study found association between *SLC9A3* and atopy and asthma [26], whereas SNPs in *SLC9A3* were also associated with lung function in cystic fibrosis patients [27, 28]. These data provide further support for our prior hypothesis [29] that lung immune responses associate with protection from TB such as previously observed in patients with asthma [30, 31]. In further support of this hypothesis, *SLC9A3* is also a component of a biomarker that predicts progression to TB [32]. A recent GWAS of lung function identified an association with *CEP72* [33], which is also in this region with associated methylation findings, so it possible that either, or both, gene(s) in this region are involved in TB susceptibility.

The results from our connected analyses (MWAS, genetic association, meQTL, and methylation x SNP interaction) are challenging to interpret, but do lay out a possible connection between genetic variation, its implications for methylation/gene regulation and TB. Nominally significant SNP association, meQTLs, and/or SNP x methylation interaction findings co-localize in regions that map to genes of interest, though different SNPs were associated in each of these terms. One explanation is because linkage disequilibrium patterns vary across African populations and the true functional variants are tagged differently across cohorts [34]. Limited sample size, combined with variable allele frequencies and distributions of the methylation marker, are additional factors. A second potential explanation for non-exact replication across cohorts may be exposure to different Mycobacterial lineages and/or environmental differences, such as cooking method, that may affect cellular phenotype. This study does not distinguish between methylation differences induced by Mtb stimulation, which has been shown by other studies [17, 19, 35, 36] versus inherent differences between individuals with TB or LTBI. An alternative hypothesis may be that subjects who resist development of TB respond to Mtb-induced methylation differently; future studies are needed to distinguish these hypotheses. Larger sample sizes will be important to elucidate the effects of each of these factors. Nonetheless, our results taken as a whole indicate that methylation and genetic variation are both important factors in TB susceptibility, and the effects are not necessarily independent of each other.

It is important to emphasize that the study subjects in this analysis were not on ART or anti-TB treatment at the time that blood was drawn for this study, as methylation profiles may be influenced by ART [37-39]. This is a strength of our study although given the ubiquity of ART, future replication studies will only identify differentially methylated regions that are robust in patients on ART. Another potential limitation of our study is the use of a broad array instead of bisulfite sequencing, which examines only select nucleotides for epigenetic modification. HIV may have an impact on differential methylation, but since all subjects in this study were HIV-infected, that potential confounder was controlled; this may result in findings that are not generalizable to HIV-uninfected. Our MWAS results do not attain multiple testing correction, but the support of the aforementioned SNP association tests bolster confidence in these findings. Lastly, the source of DNA for this assay was buffy coat, which consists of a variety of cell types, including the major cells involved in the TB immune response, potentially enriching findings significant to those cell types compared to others. Nonetheless, these findings clearly indicate future studies should explore the linkage between epigenetic regulation of cellular and humoral immune responses with protection from TB along with other genomic data.

It is also of interest to understand how differential methylation affects RNA expression. However, this question in TB is not trivial. The most easily accessible tissue, peripheral blood mononuclear cells, may not be the relevant tissue for TB. Some TB transcriptomic studies [40] stimulate monocyte-derived macrophages and examine the RNA expression change after stimulation with Mtb. While this gets closer to the immune response, it is more difficult to measure than RNA expression level at a putative baseline that is usually what is studied [12]. The primary cell of interest would be alveolar macrophages. There are ongoing studies to determine whether gene expression in the lung is different than in circulating blood.

## CONCLUSION

In conclusion, this is the first study to identify methylation changes associated with protection from active TB in patients with HIV-associated susceptibility to TB disease. Methylation is of interest because it is a possible pharmacologic target that describes cellular response to MTB infection and disease progression. Future studies will need to extend our findings and examine the impact of methylation on differential RNA expression and how these can vary by patterns of genetic variation, especially in regulatory regions, and how methylation profiles differ by specific cell type. However, it is now clear that patterns of methylation and genetic variation synergize with each other and can identify important associations in TB risk.

## METHODS

### Study participants

This study includes the same subjects as in our previous GWAS [5]. Briefly, subjects were from a household contact study in Kampala, Uganda [41], or two clinical trials of TB in Dar es Salaam, Tanzania [42]. All subjects were HIV-infected, over the age of 15, and none received ART due to unavailability at the time of enrollment. TB cases were culture-confirmed. Controls all had LTBI based on a positive tuberculin skin test; interferon-γ release assays were not performed at the time of subject ascertainment. All subjects provided written informed consent. The present analyses utilized samples from those subjects who had DNA remaining after the GWAS.

### Molecular methods for MWAS and GWAS

DNA came from buffy coat samples in the Uganda cohort and either buffy coat or whole blood in the Tanzania cohort, and was prepared as previously described [5] then bisulfite converted according to the specifications for the Illumina Methylation EPIC 850k chip. GWAS data were available from our previous analysis [5].

### Quality control and statistical analysis

#### Quality control and principal components analysis

Quality control and normalization of raw methylation data (as Illumina .idat files) were applied to the combined Uganda and Tanzania cohorts and carried out using the Bioconductor package minfi for R [43, 44]. Data for probes and samples with good detection p-values (less than 0.01) were retained. We normalized signal intensity by means of the BMIQ algorithm [45], which adjusts for differences between Infinium I and II probes, adjusted for batch effects using ComBat [46] and adjusted for between-array technical variation, as well as, background variation using the functional normalization procedure [47].

We estimated cell proportions in the combined Uganda and Tanzania cohort for CD8, CD4, natural killer, B, monocyte and neutrophil cells using the minfi package. Within each cohort, we conducted a methylome-wide principal components analysis, using the prcomp function in R. We regressed our TB outcome on each of the first 20 principal components (PCs) selecting PCs that were significantly associated (at p-value < 0.1) with TB in a model that included cell proportions. This resulted in multiple PCs and cell proportions (dependent on cohort) retained as covariates in each cohort specific MWAS.

#### MWAS Analysis

Within each of the two cohorts we tested for association between CpG beta values (converted to M values (log2(β/1-β)) and TB status using limma in R [48]. Here we adjusted for significant methylation-based PCs, cell proportions, age and sex in a linear model that compares methylation values between TB and LTBI samples. Because Uganda was the discovery cohort, we sought to replicate top hits from the Uganda cohort (at p-value < 5 ×10^−5)^ in the Tanzania cohort (with p-value < 0.10).

#### Genetic association analysis

Genetic association analysis was performed using the full data from Sobota et al. [5] Association between SNPs in the regions of interest and TB was conducted using PLINK [49] for SNPs with minor allele frequency > 0.10. We examined SNPs within ± 200 kb of the associated methylation marker. A Bonferonni-corrected p-value was derived based on the total number of SNPs tested across all 3 regions (20 SNPs on chromosome 1, 33 on chromosome 2, and 22 on chromosome 3).

#### meQTL analysis

We conducted a targeted cis-methylation QTL analysis around the replicated CpG sites in the combined Uganda/Tanzania sample that had overlapping methylation and genotype data (total N = 188, with 75 from the Tanzania cohort (32 with TB) and 113 from the Uganda cohort (66 with TB)). SNPs within ±200 kb of the replicated CpG sites were pulled from the GWAS dataset [5]. Using Matrix eQTL, we ran a linear regression model adjusting for gender, age, cohort, cell proportions (CD4, CD8, monocytes, neutrophils) and the first two genetic-based PCs [5] to determine if nearby SNPs were influencing the methylation levels of our replicated CpG sites.

#### SNP x methylation interaction

We conducted a targeted interaction analysis around the replicated CpG sites in the combined Uganda/Tanzania sample that had overlapping methylation and genotype data (sample sizes as above). As in the meQTL analysis, SNPs within 200 kb of the replicated CpG sites were pulled from the GWAS dataset [5]. Using glm in R we ran a logistic regression model adjusting for gender, cohort, CD4 cell proportions and the first two GWAS-based PCs [5] to determine if the difference in methylation levels between TB and LTBI samples are modified by genotypes of nearby SNPs.

### Functional annotation

HUGIn [50] and RegulomeDB [51] were used to examine chromatin state evidence predicting whether or not our methylation markers fell in promoter or enhancer regions, whether associated methylation markers were in regions of DNAase hypersensitivity, transcription factor biding sites, and promoter regions, and also to identify frequently interacting regions (FIREs).

## Data Availability

Summary data (marker names and p-values) will be provided at Dryad (https://github.com/CDL-Dryad/dryad). The Ugandan IRB did not grant approval for broad genetic data sharing, so individual-level data may be requested from the Data Access Committee, chaired by Dr. Sudha Iyengar. Methylation data from the Tanzanian cohort will be available at Dryad (https://github.com/CDL-Dryad/dryad).

## LIST OF ABBREVIATIONS

ARVs: anti-retroviral therapy
GWAS: genome-wide association study
HIV: Human Immunodeficiency virus
LTBI: latent *M. tuberculosis* infection
meQTL: methylation quantitative trait locus
Mtb: *Mycobacterium tuberculosis*
MWAS: methylome-wide association study
SNP: single nucleotide polymorphism
TB: Tuberculosis

## DECLARATIONS

### Ethics approval and consent to participate

Informed consent was obtained from all patients in the extended DarDar follow-up cohort. The research ethics committee at the Muhimbili University of Health and Allied Sciences and the Committee for the Protection of Human Subjects at Dartmouth College and the Dartmouth-Hitchcock Medical Center approved this study. Informed consent was obtained from all subjects in the Household Contact study in Kampala, Uganda. Ethics committees that approved this work were at Muhimbili University of Health and Allied Sciences, Committee for the Protection of Human Subjects at Dartmouth College (#14606), Uganda Council for Science and Technology, and University Hospitals of Cleveland (10-01-25)

### Consent for publication

N/A

### Authors’ contributions

CMS, SMW, and GS conceived of the study. CS, SMW, CM, WSB, and WKS directed the statistical analyses. WKS supervised genotyping and methylation typing. PB, JB, RSS, and RPI, Jr conducted the statistical analyses. CMS, H M-K, WHB, CFvR, and TL oversaw the clinical characterization of the subjects. KC and RSS obtained DNA samples and supervised shipment of samples to the laboratory. CMS and PB drafted the initial manuscript. All authors read and approved the final manuscript.

## Acknowledgments

This work made use of the High Performance Computing Resource in the Core Facility for Advanced Research Computing at Case Western Reserve University. We would like to acknowledge the invaluable contribution made by the study medical officers, health visitors, laboratory and data personnel: Dr. Lorna Nshuti, Dr. Roy Mugerwa, Dr. Sarah Zalwango, Allan Chiunda, Bonnie Thiel, Mark Breda, Dennis Dobbs, Hussein Kisingo, Mary Rutaro, Albert Muganda, Richard Bamuhimbisa, Yusuf Mulumba, Deborah Nsamba, Barbara Kyeyune, Faith Kintu, Dr. Mary Nsereko, Gladys Mpalanyi, Janet Mukose, Grace Tumusiime, Pierre Peters, Annet Kawuma, Saidah Menya, Joan Nassuna, Alphonse Okwera, LaShaunda Malone, Keith Chervenak, Karen Morgan, Alfred Etwom, Micheal Angel Mugerwa, Dr. Brenda Okware, and Lisa Kucharski. We would like to acknowledge and thank Dr. Francis Adatu Engwau, Head of the Uganda National Tuberculosis and Leprosy Program, for his support of this project. We would like to acknowledge the medical officers, nurses and counselors at the National Tuberculosis Treatment Centre, Mulago Hospital, the Ugandan National Tuberculosis and Leprosy Program and the Uganda Tuberculosis Investigation Bacteriological Unit, Wandegeya, for their contributions to this study. We would also like to thank the staff at the Infectious Disease Center in Dar es Salaam, Tanzania, and Betty Mchaki and Mecky Matee at Muhimbili University of Health and Allied Sciences. This study would not be possible without the generous participation of the Ugandan and Tanzanian patients and families.

## Supplemental Tables

**Supplemental Table 1:** SNP association results within regions identified through MWAS.

**Supplemental Table 2**: Follow-up of regions identified in Sobota et al. [5] within MWAS data. MWAS p-values are provided separately for Ugandan and Tanzanian cohorts, and combined p-values are indicated as (*p-value) within the cells with both Ugandan and Tanzanian cohort p-values.

